# Willingness to Pay for Primary Health Care Services and Associated Factors in Eastern Kasai, Democratic Republic of the Congo

**DOI:** 10.64898/2026.05.21.26353764

**Authors:** Barry Mutombo, Alain Cimuanga-Mukanya, Pascal Lutumba

## Abstract

**Background:** In the Democratic Republic of the Congo (DRC), health care financing relies heavily on out-of-pocket payments, limiting access to essential services. In a context of declining external funding and ongoing efforts toward Universal Health Coverage (UHC), understanding households’ willingness to pay (WTP) for health care is critical for designing sustainable financing strategies. This study aimed to assess WTP for primary health care services and identify its associated factors in Eastern Kasai Province.

**Methods:** A cross-sectional study based on the contingent valuation method was conducted from 10 to 30 July 2025 among 633 randomly selected households using a multistage probabilistic sampling approach. Data were collected through semi-structured interviews using KoboToolBox. WTP was assessed using a stated preference approach. Logistic regression analyses using R 4.5.0 were performed to identify factors associated with WTP at a significance level of p < 0.05. Adjusted odds ratios (aORs) with 95% confidence intervals (95% CI) were reported.

**Results:** Overall, 70% of household heads reported willingness to pay for their own health care, and 73% for other household members. WTP decreased significantly as the cost of services increased, dropping from 95.5% for free care to 6.3% at the highest cost levels (above CDF 230,000). Poor perceived quality of care was a consistent reason for refusal, alongside financial constraints such as low income and indebtedness. Multivariable analysis showed that having a professional activity (OR = 1.9; 95% CI: 1.2–3.0; p = 0.006), residence in rural areas (OR = 2.1; 95% CI: 1.3–3.7; p = 0.008), and higher household income (OR = 2.2; 95% CI: 1.2–4.0; p = 0.011) were significantly associated with WTP. Despite relatively low absolute health care costs, the majority of households perceived them as high.

**Conclusion:** Willingness to pay for health care services in Eastern Kasai is moderate but highly sensitive to cost and strongly influenced by socioeconomic conditions and perceived quality of care. These findings underscore the need to strengthen financial protection mechanisms, particularly prepayment and risk-pooling systems, while improving service quality to enhance health care utilization and progress toward UHC in the DRC.

## 1. Introduction

The Democratic Republic of the Congo (DRC) remains one of the poorest and most economically fragile countries both in Africa and worldwide. In 2024, its gross domestic product (GDP) per capita was estimated at US$722, reflecting a 7% increase compared with 2023 and a 32% rise relative to 2014, indicating sustained economic growth over the past decade [1]. Nevertheless, these improvements have not translated into substantial gains in population living standards, which remain significantly below the global average [2], thereby constraining access to quality health care services.

In response to these financial barriers, the Congolese government has set an ambitious objective for its health system: achieving Universal Health Coverage (UHC). The implementation of this vision was initiated in July 2024 through the rollout of a free maternity and neonatal care program, with plans to extend coverage to all 26 provinces by 2026. Ensuring the sustainability of this reform requires strengthening two key pillars: the mobilization of domestic resources and increased public investment in the health sector. However, these efforts are challenged by the historically low prioritization of health within national budget allocations, as highlighted by the World Health Organization (WHO) in 2021 [3].

Health financing in the DRC remains heavily reliant on households, which contribute between 40% and 43% of total health expenditures, alongside external funding sources. Between 2019 and 2023, per capita health expenditure was estimated at approximately USD 22 – 23 [4]. Out-of-pocket payments constitute the dominant mechanism of household financing, representing 91% – 92% of their current health expenditures through payments made at health facilities. In contrast, prepayment mechanisms – whether mandatory or voluntary – remain limited at 7%, while social contributions account for only 1% [4]. In Eastern Kasai, household contributions may be even higher, reaching up to 70% of total health expenditures, with an estimated annual per capita health spending of USD 29 (Mutombo Barry et al., unpublished data, 2025). Although national projections aim to increase per capita health expenditure to USD 64 by 2030 [5], this target remains below international recommendations, which range from USD 74 to USD 198 for lower-middle-income countries [6]. Consequently, households remain the primary financiers of the health system, despite limited evidence regarding their willingness to pay (WTP) for health care services.

Across sub-Saharan Africa, WTP for health services varies considerably between settings and is influenced by multiple socio-demographic and economic factors. In South Africa, WTP has been estimated to range from 35% to 60%, depending on the type of service – such as primary health care or insurance schemes – and is associated with employment status, income level, and perceived quality of health infrastructure [7], as well as marital status and household size [8]. Other determinants include age, sex, educational attainment, health status, treatment preferences, type of care (outpatient or inpatient), perceptions of health financing, distance to health facilities, travel time, and the urban–rural context [8]. In The Gambia, WTP for the national health insurance scheme reached 94.4% and was negatively associated with sex, education level, and income [9]. In Ethiopia, reported WTP ranged from 35.4% to 42.3% [10–11], with associated factors including the presence of children aged 6 – 18 years in the household, prior experiences of unaffordable health expenditures within the past 12 months, and awareness of the social health insurance program [11]. Similarly, high levels of WTP have been reported in Nigeria and Sierra Leone (86.6% and 94.3%, respectively), primarily associated with education level, sex, and place of residence [12–13]. A multicountry study conducted in Kenya, Uganda, Nigeria, and Tanzania reported an overall WTP of 78.8%, with key associated factors including age, sex, marital status, occupation, enrollment in other health insurance schemes, monthly health expenditures, and education level [14].

In 2025, several African countries experienced a notable decline in Official Development Assistance (ODA), estimated between 9% and 17%, alongside growing uncertainty regarding future funding trends, particularly contributions from the United States of America. In this context – marked by potential reductions in external funding and the continued reliance of the DRC health system on such resources – it becomes essential to assess the value that populations assign to health care services through their willingness to pay, based on stated preferences [15–16]. Accordingly, this study aims to examine willingness to pay for health care services and its associated factors in Eastern Kasai Province, DRC.

## 2. Methods

### 2.1. Study setting

Eastern Kasai Province is located near the geographical center of the Democratic Republic of the DRC and is the smallest among the country’s 26 provinces, covering an area of 9,545 km². In 2025, its population was estimated at 6,186,353 inhabitants, corresponding to a density of approximately 629 inhabitants per km². The provincial capital, Mbujimayi, occupies less than 1% of the total land area but accommodates over 60% of the province’s population. Administratively, the province is divided into five territories and five urban municipalities, as illustrated in **Figure 1**.

**Figure 1:**
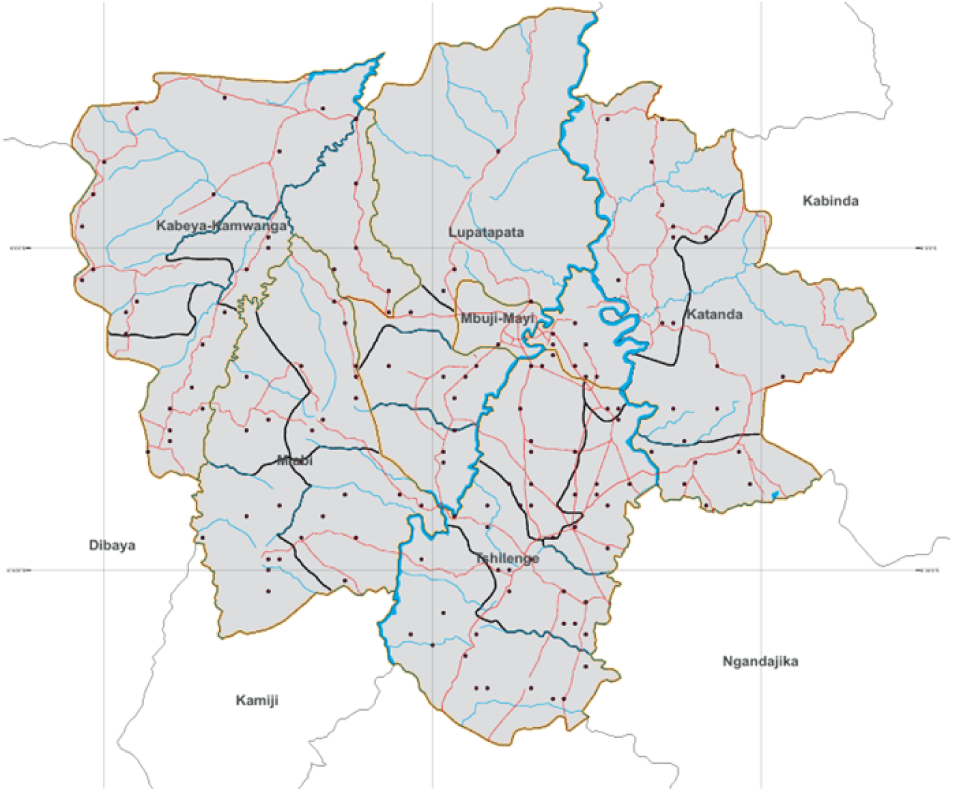
Political and administrative map of Eastern Kasai province.

The health system is structured into 19 health zones, including 10 urban and 9 rural zones, as detailed in **Table 1**. These are further subdivided into 317 health areas, themselves organized into 1,872 community outreach cells to facilitate proximity-based service delivery.

**Table 1:**
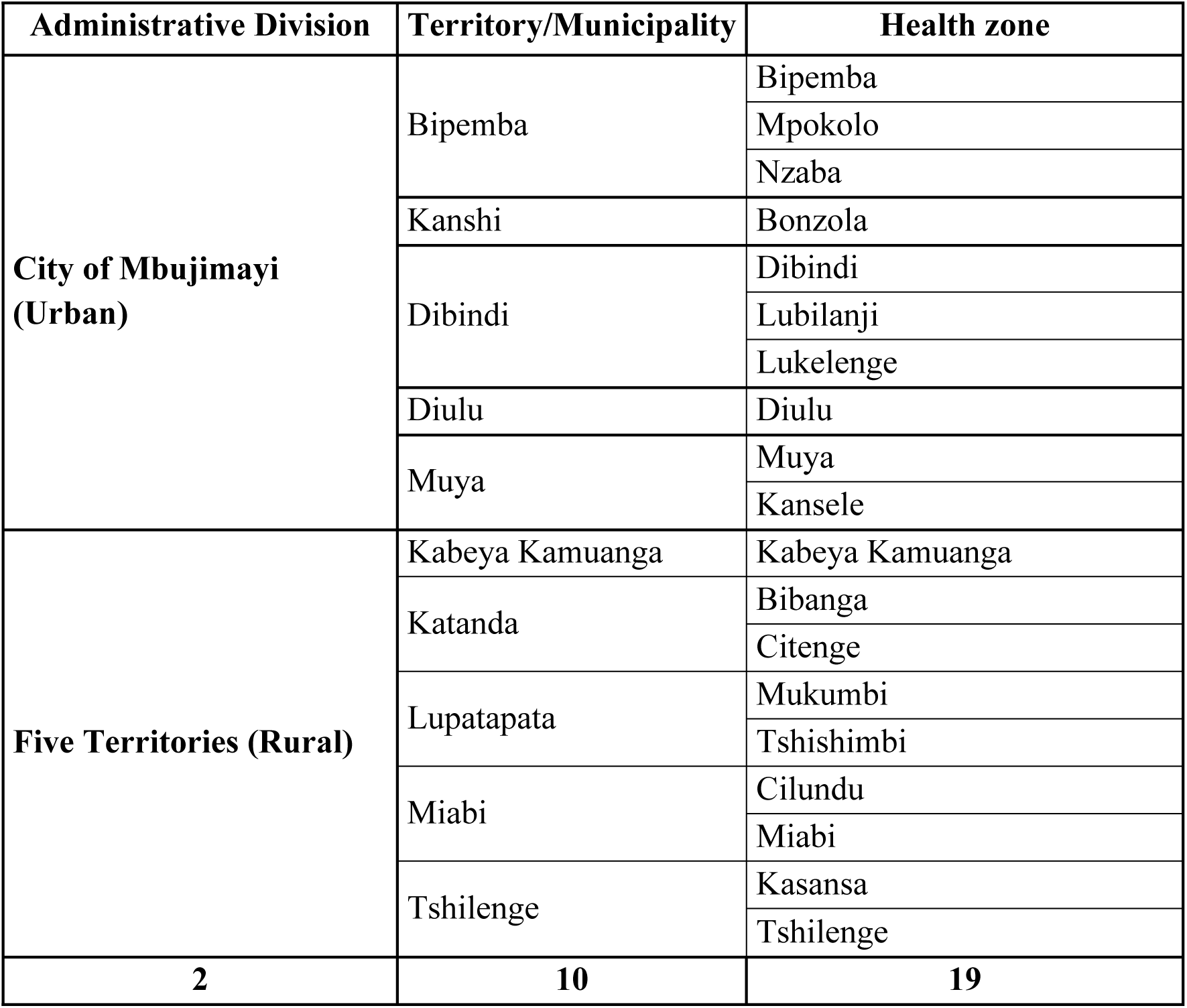
Administrative and health zone subdivision of Eastern Kasai province.

| Administrative Division | Territory/Municipality | Health zone |
| --- | --- | --- |
| **City of Mbuji-Mayi (Urban)** | Bipemba | Bipemba |
|  |  | Mpokolo |
|  |  | Nzaba |
|  | Kanshi | Bonzola |
|  | Dibindi | Dibindi |
|  |  | Lubilanji |
|  |  | Lukelenge |
|  | Diulu | Diulu |
|  | Muya | Muya |
|  |  | Kansele |
| **Five Territories (Rural)** | Kabeya Kamuanga | Kabeya Kamuanga |
|  | Katanda | Bibanga |
|  |  | Citenge |
|  | Lupatapata | Mukumbi |
|  |  | Tshishimbi |
|  | Miabi | Cilundu |
|  |  | Miabi |
|  | Tshilenge | Kasansa |
|  |  | Tshilenge |
| **2** | **10** | **19** |

Despite this organization, the province faces considerable economic challenges, with a monetary poverty rate estimated at 81.3% [17].

### 2.2. Study design and period

This study employed a cross-sectional observational design based on the contingent valuation method. Data were collected between 10 and 30 July 2025 in Eastern Kasai Province, Democratic Republic of the Congo.

### 2.3. Study population and sampling

The study population consisted of household heads residing in selected health zones. Household heads were chosen due to their central role in decision-making regarding health-related expenditures.

A multistage probabilistic sampling approach (five-stage sampling) was used. The selection process included: (i) health zones, (ii) health areas within selected zones, (iii) villages and/or neighborhoods, (iv) households, and (v) the respondent (household head).

The sample size of 633 households was determined using the StatCalc module of Epi Info software, based on a 95% confidence level, a 5% margin of error, and a design effect of 1.5, in line with parameters used in the 2018 MICS Malaria Survey in the DRC [18]. The calculation also accounted for an estimated population of 6,186,353 inhabitants and a 10% non-response rate.

Households were allocated proportionally across selected health zones according to their demographic weight (Table 2).

**Table 2:** Distribution of surveyed households across selected health zones.

| HZ | Location | Population | Number of Households |  | Expected Number of Households |
| --- | --- | --- | --- | --- | --- |
|  |  |  | Proportion | % |  |
| DIULU | Urban | 496 849 | 93 745 | 18% | 117 |
| BONZOLA | Urban | 240 596 | 45 396 | 9% | 56 |
| DIBINDI | Urban | 360 103 | 67 944 | 13% | 85 |
| MUYA | Urban | 446 510 | 84 247 | 17% | 105 |
| BIPEMBA | Urban | 204 250 | 38 538 | 8% | 48 |
| CITENGE | Rural | 318 584 | 60 110 | 12% | 75 |
| TSHILENGE | Rural | 388 506 | 73 303 | 14% | 91 |
| TSHISHIMBI | Rural | 241 723 | 45 608 | 9% | 57 |
| Total |  | 2 697 121 | 508 891 | 100% | 633 |

### 2.4. Selection criteria

Eligible participants were household heads aged 18 years or older, regardless of sex, who provided informed consent. In polygamous households, only one household unit was included. Individuals unable to respond to the questionnaire due to severe illness or cognitive impairment were excluded.

### 2.5. Variables

Two categories of variables were considered: dependent and independent variables (**Supplementary Table 3**).

For respondents willing to pay, the maximum amount they were willing to contribute and the upper threshold they would not exceed were recorded. Those unwilling to pay were asked to provide reasons for refusal.

The proposed payment ranges were based on the current cost of primary health care services, the estimated annual per capita health expenditure in Eastern Kasai (USD 29; 95% CI: 23 – 37) (Mutombo Barry et al., unpublished data, 2025), and international recommendations for lower-middle-income countries (USD 74 – 198 per capita per year) [6], expressed in local currency.

The exchange rate used was approximately 2,290 Congolese francs (CDF) per USD, based on Central Bank data for December 2025.

### 2.6. Data collection

Data were collected using a semi-structured interview questionnaire developed and digitized on the KoboToolBox platform and administered via the KoboCollect mobile application on Android smartphones. Data were transmitted in real time to a centralized server.

Prior to data collection, the questionnaire was pretested to ensure clarity, relevance, and internal consistency. Necessary adjustments were made accordingly. Enumerators received training on data collection procedures, interview techniques, sampling methodology, and use of digital tools.

### 2.7. Data analysis

Data were exported to Microsoft Excel (365), cleaned, and analyzed using R software version 4.5.0 (R Core Team, 2025). Categorical variables were summarized using frequencies and proportions, while continuous variables were described using medians, interquartile ranges (IQR), and extreme values. Willingness to pay (WTP) was assessed using the stated preference approach [15–16]. Factors associated with WTP were identified through logistic regression. Variables with p < 0.25 in bivariate analysis were included in a multivariable model based on the Generalized Linear Model framework [19]. Statistical significance was set at p < 0.05, and adjusted odds ratios (aORs) were reported with 95% confidence intervals.

### 2.8. Ethical considerations

The study adhered to the principles of the Declaration of Helsinki. Ethical approval was obtained from the Ethics Committee of the University of Mbujimayi (UM) (approval number: 02/CEI/UM/ESU/2025). Informed consent was obtained from all participants, and strict confidentiality of data was ensured. Data were stored in secured Excel files accessible only to the research team.

## 3. Results

A total of 633 households were included in the study, distributed across eight health zones (five urban and three rural).

### 3.1. Sociodemographic and economic characteristics

Household heads were predominantly aged between 30 and 49 years, with a median age of 43 years (range: 19–89). Women represented the majority (57.2%), and most respondents were married (87.7%). The main occupations were trading (36.4%) and farming (22.4%). Primary education was the most common level attained (47.2%), and more than half of respondents reported affiliation with revivalistic churches (53.2%). Participants were mainly drawn from the Diulu (18.5%) and Muya (16.6%) health zones. Household size most frequently ranged from 6 to 9 members (46%), with a median of 6 persons (range: 1–30). The most common monthly income category, both for household heads and households, ranged between 164,266 and 1,000,000 CDF (52.3% and 60.8%, respectively) (**Table 4)**.

**Table 3:** Reasons for refusal to pay for health care among household heads by cost range (CDF)

| Cost range<br>(CDF) | Household<br>heads (n) | Reasons for refusal to pay |  |  |  |  |  |  |
| --- | --- | --- | --- | --- | --- | --- | --- | --- |
|  |  | Lack of<br>income/debt<br>(%) | Low<br>income<br>(%) | Costs<br>too<br>high<br>(%) | Unemployment<br>(%) | quality<br>of care<br>(%) | Household<br>burden<br>(%) | Other<br>(%) |
| 0 | 2 | - | - | - | - | 100 | - | - |
| 1 000 - 2 500 | 9 | 44,4 | - | - | 22,2 | 22,2 | - | 11,2 |
| 2501 - 7 500 | 45 | 66,6 | 15,6 | - | 6,7 | 4,4 | - | 6,7 |
| 7 501 - 15 000 | 101 | 68,3 | 17,8 | 8,9 | 4 | 1 | - | - |
| 15 001 - 29 000 | 183 | 68,8 | 12,6 | 14,2 | 3,3 | - | - | 1,1 |
| 29 001 - 58 000 | 253 | 64 | 14,6 | 17,8 | 2,4 | - | 1,2 | - |
| 58 001 - 107 000 | 329 | 57,7 | 18,5 | 19,5 | 3,3 | - | 0,9 | 0,1 |
| 107 001 - 185 000 | 381 | 57,3 | 23,1 | 17,6 | 1,6 | - | 0,5 | - |
| 185 001 - 231 000 | 412 | 53,8 | 26,9 | 17,7 | 1,2 | - | 0,2 | 0,2 |
| 231 001 - 571 000 | 432 | 51,6 | 27,5 | 18,1 | 1,6 | - | 1,2 | - |

**Table 4:**
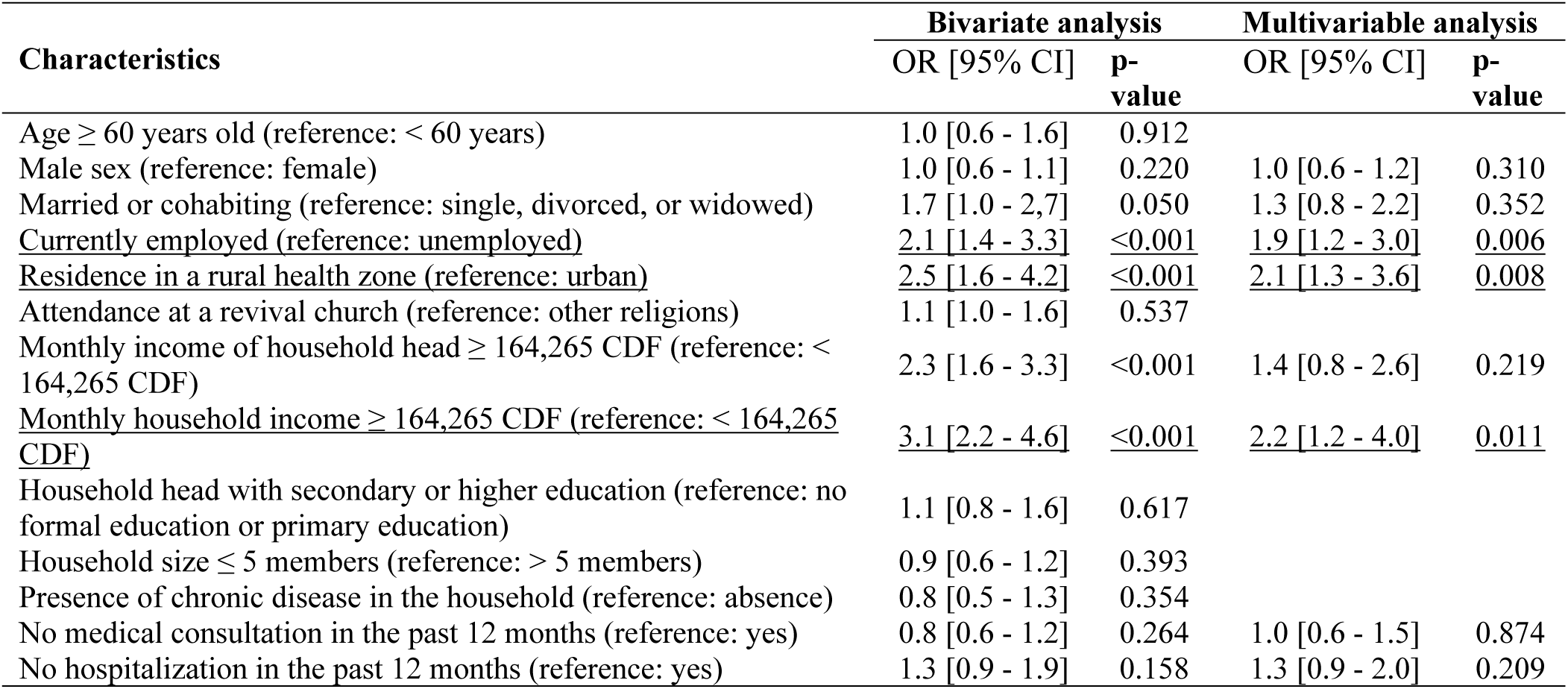
Factors associated with willingness to pay for health care services.

| Characteristics | Bivariate analysis |  | Multivariable analysis |  |
| --- | --- | --- | --- | --- |
|  | OR [95% CI] | p-value | OR [95% CI] | p-value |
| Age ≥ 60 years old (reference: < 60 years) | 1.0 [0.6 - 1.6] | 0.912 |  |  |
| Male sex (reference: female) | 1.0 [0.6 - 1.1] | 0.220 | 1.0 [0.6 - 1.2] | 0.310 |
| Married or cohabiting (reference: single, divorced, or widowed) | 1.7 [1.0 - 2,7] | 0.050 | 1.3 [0.8 - 2.2] | 0.352 |
| Currently employed (reference: unemployed) | 2.1 [1.4 - 3.3] | <0.001 | 1.9 [1.2 - 3.0] | 0.006 |
| Residence in a rural health zone (reference: urban) | 2.5 [1.6 - 4.2] | <0.001 | 2.1 [1.3 - 3.6] | 0.008 |
| Attendance at a revival church (reference: other religions) | 1.1 [1.0 - 1.6] | 0.537 |  |  |
| Monthly income of household head ≥ 164,265 CDF (reference: < 164,265 CDF) | 2.3 [1.6 - 3.3] | <0.001 | 1.4 [0.8 - 2.6] | 0.219 |
| Monthly household income ≥ 164,265 CDF (reference: < 164,265 CDF) | 3.1 [2.2 - 4.6] | <0.001 | 2.2 [1.2 - 4.0] | 0.011 |
| Household head with secondary or higher education (reference: no formal education or primary education) | 1.1 [0.8 - 1.6] | 0.617 |  |  |
| Household size ≤ 5 members (reference: > 5 members) | 0.9 [0.6 - 1.2] | 0.393 |  |  |
| Presence of chronic disease in the household (reference: absence) | 0.8 [0.5 - 1.3] | 0.354 |  |  |
| No medical consultation in the past 12 months (reference: yes) | 0.8 [0.6 - 1.2] | 0.264 | 1.0 [0.6 - 1.5] | 0.874 |
| No hospitalization in the past 12 months (reference: yes) | 1.3 [0.9 - 1.9] | 0.158 | 1.3 [0.9 - 2.0] | 0.209 |

### 3.2. Clinical characteristics of households

Overall, 21.5% of household heads reported the presence of at least one chronic condition within their household (**Figure 2A**). Among these, hypertension was the most commonly reported (43.4%), followed by tuberculosis (14%), gastritis and/or gastroduodenal ulcer (12.5%), hemorrhoids (11%), osteitis (10.3%), diabetes mellitus (7.4%), and hernias (5.9%) (**Figure 2B**).

**Figure 2.**
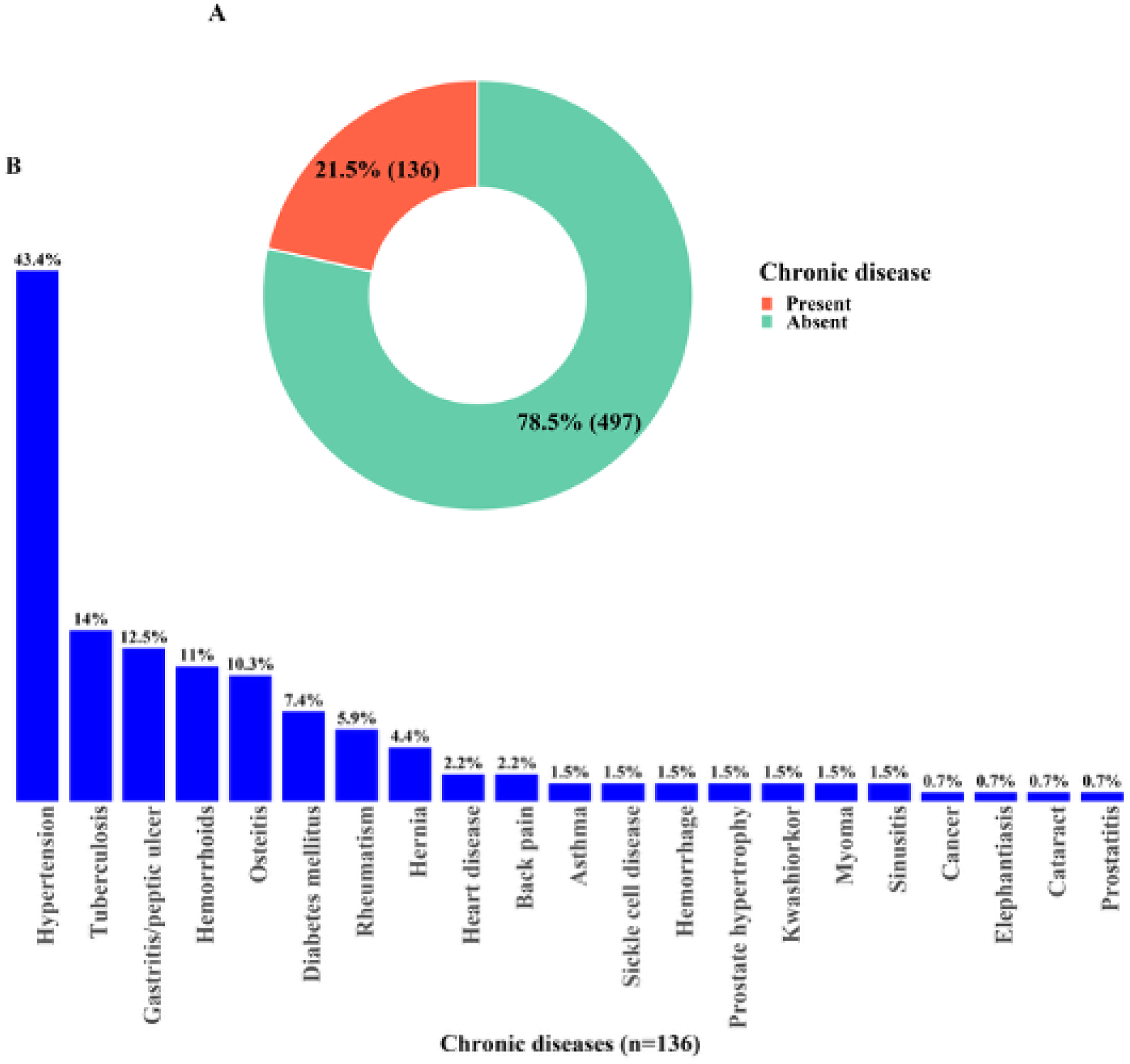
Prevalence of chronic diseases among households.

In the 12 months preceding the survey, 34% of households reported at least one medical consultation (**Figure 3A**). The majority of these consultations were related to acute conditions (86.6%), compared with 13.4% for chronic diseases (**Figure 3B**). Acute illnesses were predominantly malaria (65.7%) and typhoid fever (43.6%), followed by helminth infections (11.9%), acute gastritis (3.3%), and diarrhea (2.5%) (**Figure 3C**). Chronic conditions reported during consultations were mainly hypertension (32.1%) and heart diseases (28.6%), followed by diabetes mellitus (14.3%), tuberculosis (8.9%), and chronic gastritis (7.1%) (**Figure 3D**).

**Figure 3.**
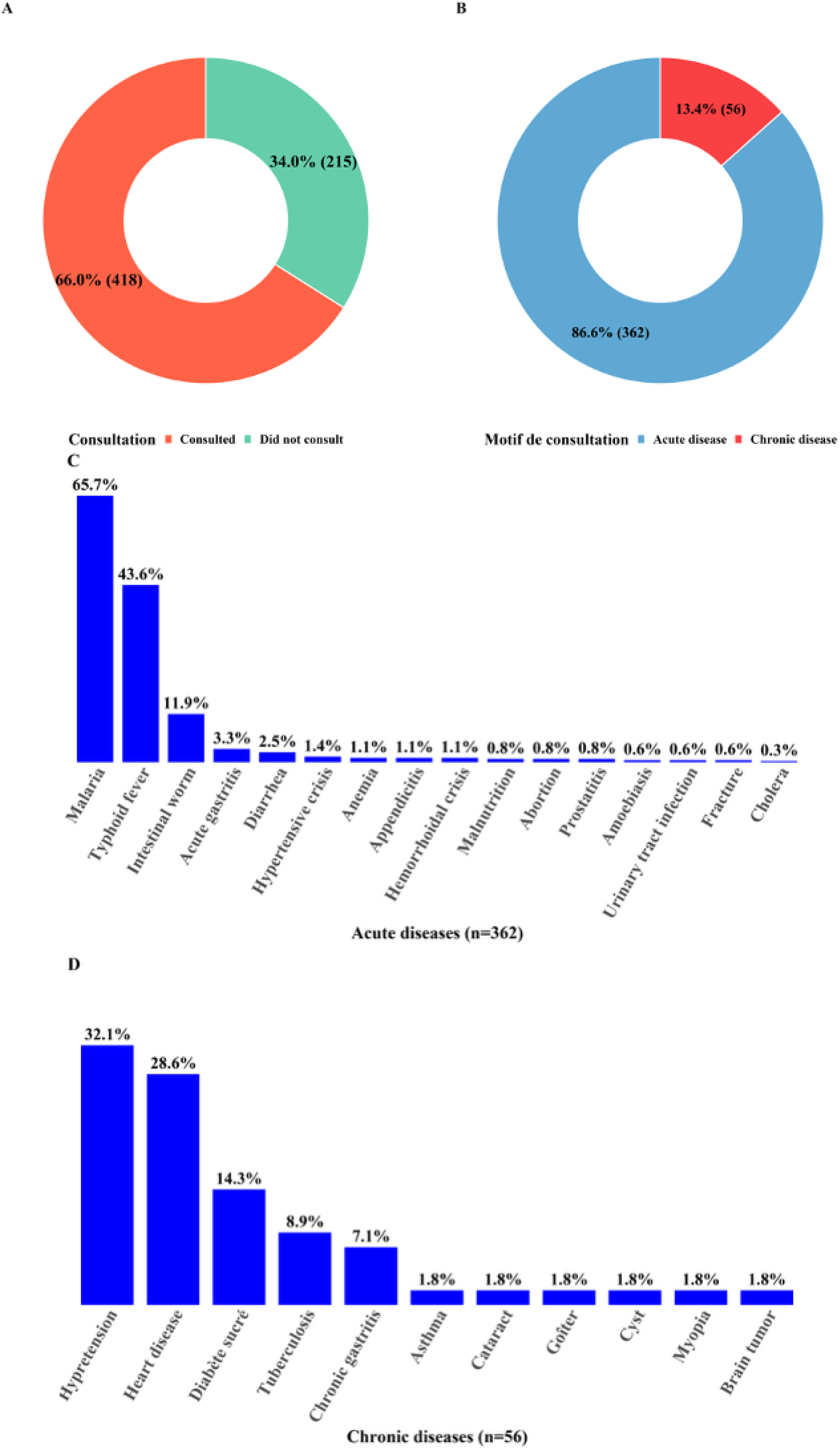
Household medical consultations in the previous 12 months and reasons for consultation.

Regarding hospitalization, 36.5% of households reported at least one hospital admission within the previous 12 months (**Figure 4A**). Most hospitalizations occurred in health centers (63.2%), while general referral hospitals accounted for 32.9% (**Figure 4B**). The leading causes of hospitalization were malaria (43.7%) and typhoid fever (33.8%), followed by anemia (8.2%), hypertension (7.4%), and surgical conditions (6.1%) (**Figure 4C**).

**Figure 4.**
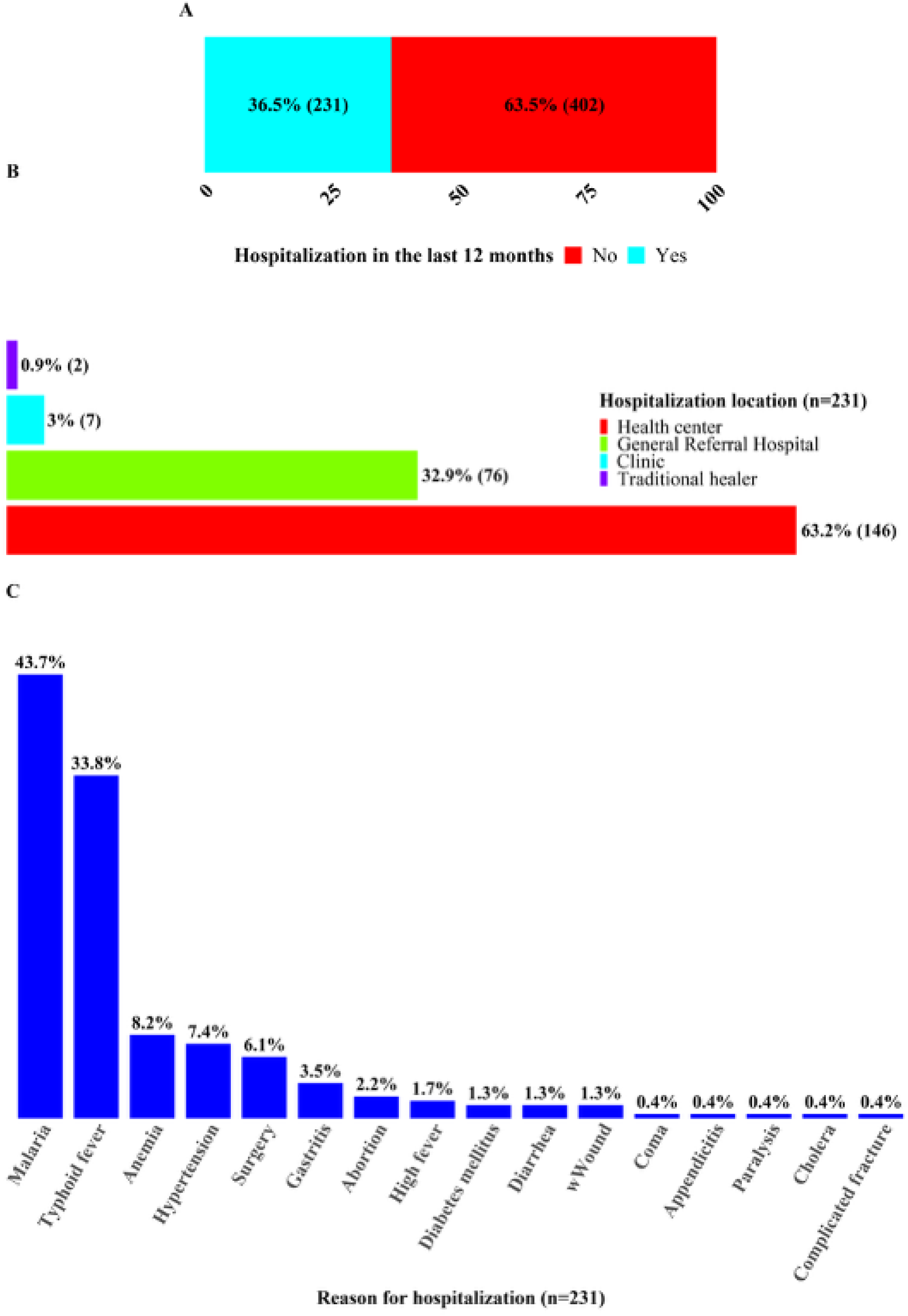
Location and reasons for hospitalization during the 12 months preceding the survey.

Hospitalization costs were below 100,000 CDF (≈43.7 USD) in 67.1% of cases, ranged between 100,000 and 200,000 CDF (≈87.3 USD) in 19% of cases, and exceeded 500,000 CDF (≈218.3 USD) in 5.6% of cases (**Figure 5A**). These costs were primarily covered by household income (60.6%), followed by external financial support (30.3%), whereas the sale of valuable assets was reported in a minority of cases (5.2%) (**Figure 5B**).

**Figure 5.**
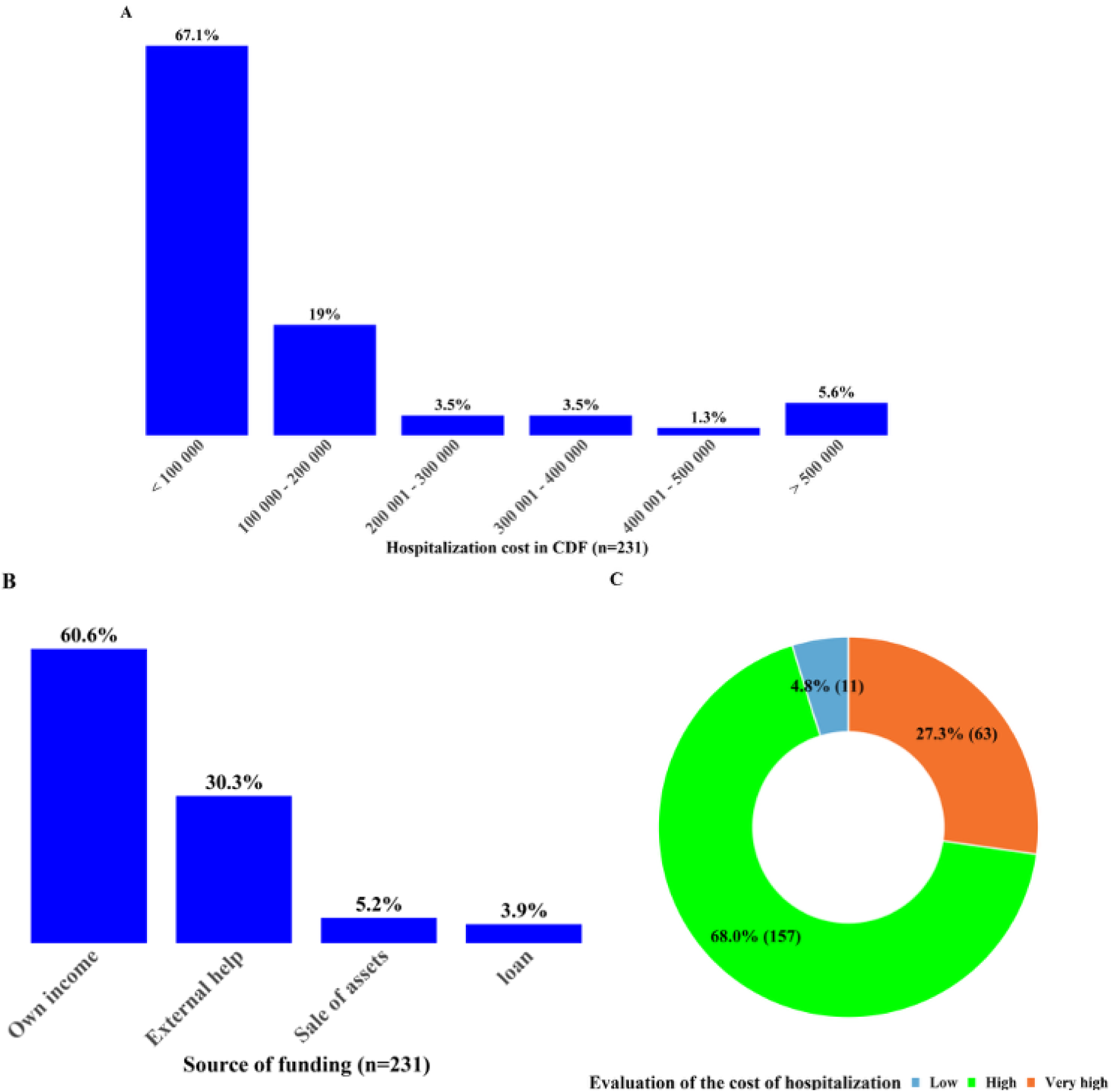
Hospitalization costs, sources of financing, and perceived cost of hospitalization.

Most household heads considered these costs high (68%) or very high (28.3%), compared with only 4.7% who regarded them as low (**Figure 5C**).

### 3.3. Willingness to pay for health care services and associated factors

The overall willingness to pay (WTP) for health care services was estimated at 70% among household heads and 73% for other household members (**Figure 6A**). However, WTP declined progressively as the cost of care increased. While nearly all respondents were willing to pay when services were free (95.5%), this proportion remained high for costs between 1,000 and 2,500 CDF (96.3%), before decreasing to 44.8% for costs ranging from 29,001 to 58,000 CDF, and further dropping to 6.3% for costs between 231,001 and 571,000 CDF. In parallel, the proportion of respondents unwilling to pay increased substantially with rising costs, reaching 93.5% in the highest cost category (**Figure 6B**).

**Figure 6.**
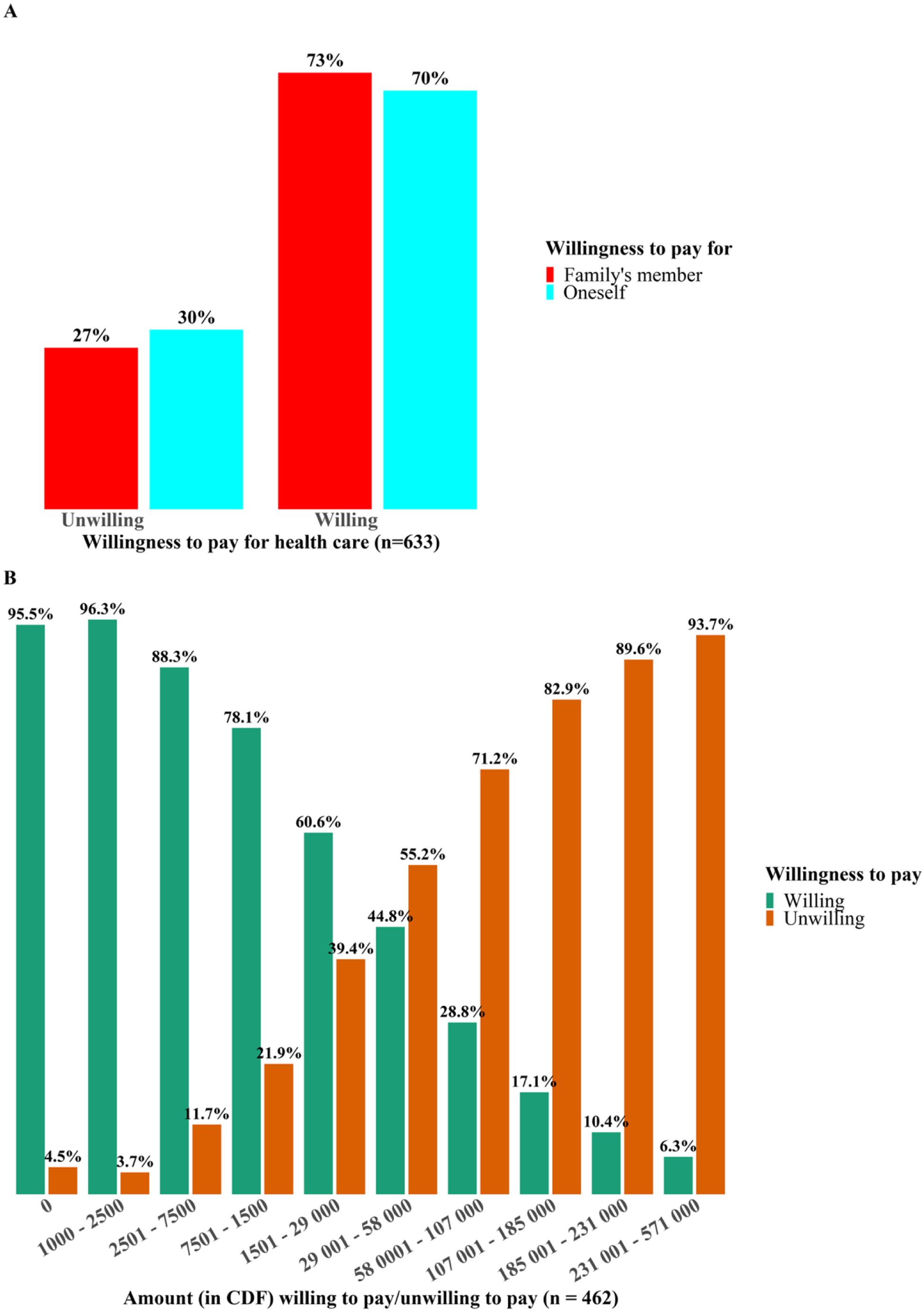
Willingness and unwillingness to pay for health care services.

Poor perceived quality of health services was consistently cited by all respondents who reported unwillingness to pay, regardless of the cost level. At lower cost ranges (1,000–7,500 CDF), the main reasons included unemployment and dissatisfaction with the quality of care. At intermediate cost levels (7,501–58,000 CDF), financial constraints – particularly low income and indebtedness – were the predominant reasons, reported in up to 68.8% of cases. At higher cost levels (>58,000 CDF), the perceived excessive cost of care emerged as the primary barrier. Household burden was only rarely mentioned (<2%) (**Table 3**).

Multivariable logistic regression analysis identified several factors significantly associated with higher willingness to pay for health care services. These included engagement in a professional activity (OR = 1.9; 95% CI: 1.2–3.0; p = 0.006), residence in a rural health zone (OR = 2.1; 95% CI: 1.3–3.7; p = 0.008), and a monthly household income ≥ 16,426.5 CDF (≈7.2 USD) (OR = 2.2; 95% CI: 1.2–4.0; p = 0.011) (**Table 4**).

## 4. Discussion

This study aimed to assess households’ WTP for health care services and to identify its associated factors in Eastern Kasai Province, Democratic Republic of the Congo. The analysis was based on a random sample of 633 households distributed across eight health zones, encompassing both urban and rural settings.

### 4.1. Sociodemographic and clinical characteristics of participants

The findings indicate that household heads were predominantly aged between 30 and 49 years, female, and married, with primary education as the most common level of schooling. This profile is broadly consistent with evidence from several studies conducted in sub-Saharan Africa, where household heads are typically within the economically active age group and share similar sociodemographic characteristics, although proportions vary across settings [20]. The predominance of primary education observed in this study is also in line with findings reported in countries such as Benin and the Gambia, although higher educational attainment has been documented in other contexts [9,13,20–21].

From a socioeconomic perspective, the predominance of trade and agriculture, coupled with a high level of informal employment, reflects the structural characteristics of the Congolese economy. This pattern – marked by employment instability and the dominance of the informal sector – has been widely reported across sub-Saharan Africa, albeit to varying degrees [9,13]. In the DRC, where informal employment accounts for nearly 80% of total employment, income instability remains a major constraint on household economic security despite ongoing macroeconomic growth [3].

Most participants resided in urban areas, particularly in the Diulu and Muya health zones, a pattern also observed in studies conducted in Sierra Leone and the Gambia [9,13]. In contrast, research from other contexts, such as Nigeria, has reported a predominance of rural populations, underscoring the role of geographical context in shaping health care access and utilization patterns [12].

The income levels observed in this study remain relatively low compared with countries such as South Africa [20], although they appear somewhat higher than those reported in Sierra Leone [13]. This variability highlights the substantial economic disparities that persist across sub-Saharan Africa. In the DRC, income instability – largely driven by dependence on the informal sector – continues to undermine household resilience [22], while in other settings, macroeconomic factors such as inflation may further erode purchasing power.

From a clinical perspective, approximately one-fifth of households reported at least one chronic condition, with hypertension being the most frequently cited. This finding is consistent with regional evidence indicating a growing burden of noncommunicable diseases, particularly hypertension, whose prevalence ranges between 16% and 40% among adults and may be higher among older populations [23–25]. The prominence of hypertension in both consultations and hospitalizations further suggests its central role in the local disease profile.

At the same time, health care utilization remains largely driven by preventable acute conditions, notably malaria and typhoid fever. This coexistence of communicable and noncommunicable diseases reflects an ongoing epidemiological transition, widely documented in sub-Saharan Africa [26]. This dual burden represents a significant challenge for health systems, which must simultaneously respond to diverse and increasing health needs.

Although hospitalization costs appear relatively low in absolute terms, they are perceived as high by the vast majority of households. This perception is likely driven by the heavy reliance on out-of-pocket payments, which constitutes a major barrier to access to care. Similar observations have been reported in Eastern Kasai, where both users and providers consider health care costs to be prohibitive [27]. More broadly, evidence from the region indicates that direct payments increase the risk of catastrophic health expenditures and household impoverishment, as shown in Kenya [28]. Consequently, out-of-pocket financing remains a key driver of inequities in access to health services [28–30].

### 4.2. Willingness to pay and associated factors

Approximately 70% of household heads reported being willing to pay for their own health care, and slightly more for other household members. These findings are broadly consistent with those reported in other African contexts, although variations exist depending on the type of financing mechanism considered, particularly between direct out-of-pocket payments and insurance-based systems [14]. In contrast to studies conducted in prepayment contexts, where WTP is generally higher, the present study was conducted in a setting dominated by direct payments, which may limit households’ effective capacity to commit financial resources to health care.

As expected, willingness to pay declined markedly with increasing health care costs, reflecting standard economic behavior related to price sensitivity [7]. Notably, however, a proportion of households reported unwillingness to pay even when services were free, citing poor perceived quality of care. This finding underscores the critical importance of service quality as a determinant of health care utilization, independently of financial considerations, as documented in previous studies [7,26,31–32].

The reasons for unwillingness to pay were primarily related to financial constraints, including low income and indebtedness, highlighting the economic vulnerability of households in this setting. These findings reflect the broader context of structural poverty in Eastern Kasai, characterized by a fragile local economy and limited income-generating opportunities.

The analysis of associated factors confirms the central role of socioeconomic conditions in shaping willingness to pay. Engagement in income-generating activities and higher household income were both significantly associated with increased WTP, in line with economic theory and empirical findings from other studies [7,14,20,33]. Interestingly, residence in rural areas was also associated with higher WTP. This may be explained by the presence of community-based solidarity mechanisms, such as informal mutual support systems, as well as by a higher perceived value of health services in contexts where supply is limited [27].

Overall, this study highlights a moderate level of willingness to pay for health care services in a context characterized by significant economic constraints and a strong reliance on out-of-pocket payments. Financial capacity emerges as a key determinant of WTP, while perceived quality of care plays a critical role in shaping health-seeking behavior.

These findings point to the need for strengthening financial protection mechanisms, particularly through the development of prepayment and risk-pooling systems, alongside efforts to improve the quality of health services, in order to enhance equitable access to care in the DRC.

This study has several limitations. The use of a contingent valuation approach based on hypothetical scenarios may introduce reporting bias, as stated preferences do not necessarily reflect actual behavior. In addition, the possibility of strategic bias cannot be excluded, as some respondents may have underestimated their willingness to pay in anticipation of future policy implications.

Despite these limitations, the study presents important strengths. It provides original evidence in a context where data remain scarce, adopts a comprehensive approach integrating sociodemographic, economic, and health dimensions, and identifies key determinants that may inform health financing policies. Furthermore, the consistency of the findings with existing literature supports their external validity.

## Data Availability

Due to ethical and confidentiality considerations associated with a household-based survey, individual-level data cannot be publicly shared. Aggregated data supporting the findings of this study are included within the manuscript and its Supporting Information files. De-identified data may be requested from the corresponding author and will be shared upon reasonable request following approval by the relevant ethics committee.

## Supplementary information

## Acknowledgements

The authors gratefully acknowledge the invaluable contribution and dedicated voluntary commitment of the students from the Department of Public Health at the University of Mbujimayi throughout the data collection process.

## Authors’ contributions

## Conflict of interest statement

The authors declare no conflicts of interest.

## Funding

This study received no external funding.

## 3.2 Factors associated with willingness to pay for health care services

## Supplementary material

**Supplementary Table 1.** Operational definition of study variables.

| Variable | Operational Definition | Scale and Categories |
| --- | --- | --- |
| **Independent Variables** |  |  |
| Sex of household head | Biological sex of the household head | Nominal:<br>• Male<br>• Female |
| Age of household head | Age in completed years at the time of the survey | Discrete quantitative |
| Marital status of household head | Marital status of the household head at the time of the survey | Nominal:<br>• Married;<br>• Single;<br>• Divorced;<br>• Widowed;<br>• Cohabiting |
| Occupation of household head | Main economic activity of the household head at the time of the survey | Nominal:<br>• Civil servant;<br>• Trader;<br>• Miner (artisanal digger);<br>• Farmer;<br>• Unemployed;<br>• Other (specify) |
| Residence of household head | Place of residence according to the health zone | Nominal:<br>• Bipemba;<br>• Diulu;<br>• Bonzola;<br>• Dibindi;<br>• Muya;<br>• Citenge;<br>• Tshilenge;<br>• Tshishimbi |
| Monthly income of household head | Monthly income earned by the household head (in Congolese francs, CDF) | Ordinal:<br><164,265;<br>164,266-1,000,000;<br>1,000,001-2,500,000; 2,500,001-4,000,000; 4,000,001-5,500,000;<br>5,500,001-7,000,000; 7,000,001-8,500,000; 8,500,001-10,000,000; 10,000,001-11,500,000; >11,500,000 |
| Religion of household head | Religion practiced by the household head | Nominal:<br>• Catholic;<br>• Protestant;<br>• Revival church;<br>• Islam;<br>• Jehovah's Witness;<br>• Baba congregation; |
|  |  | • Neo-apostolic;<br>• Branham;<br>• None;<br>• Other (specify) |
| Education level of household head | Highest level of education attained by the household head | Nominal:<br>• None;<br>• Primary;<br>• Secondary;<br>• Higher education |
| Household size | Number of individuals living in the household under the responsibility of the household head | Ordinal:<br><3 persons;<br>3-5 persons;<br>6-9 persons;<br>>9 persons |
| Monthly household income | Total monthly income of the household from all members (in CDF) | Ordinal:<br>< 164 265<br>164.266-1.000.000<br>1000.001-2.500.000<br>2.500.001-4.000.000<br>4.000.001-5.500.000<br>5.500.001-7.000.000<br>7.000.001-8.500.000<br>8.500.001-10.000.000<br>10.000.001-11.500.000<br>> 11.500.00 |
| Household assets | Valuable household assets capable of generating income | Nominal:<br>• Television;<br>• Radio;<br>• Bicycle;<br>• Mobile phone;<br>• Sewing machine;<br>• Motorcycle;<br>• Vehicle;<br>• Built plot;<br>• Unbuilt plot;<br>• Small livestock;<br>• Other |
| Presence of chronic disease in the household | Presence of at least one household member with a chronic disease (e.g., sickle cell anemia, diabetes, hypertension, cancer, etc.) | Nominal:<br>• Yes<br>• No |
| Medical consultation in the past 12 months | Whether at least one household member consulted a health facility in the previous 12 months | Nominal:<br>• Yes<br>• No |
| Reason for consultation | Type of illness leading to consultation | Nominal:<br>Acute disease<br>Chronic disease |
| Hospitalization in the past 12 months | Whether at least one household member was hospitalized in the previous 12 months | Nominal:<br>Yes<br>No |
| Cost of care paid | Amount paid for health care services (in CDF) | Ordinal:<br>≤50,000;<br>50,001-100,000;<br>100,001-150,000;<br>150,001-288,000;<br>>288,000 |
| Source of financing | Source used to pay for health care services | Nominal |
| Perception of cost of care | Perceived level of health care costs by respondents | Ordinal:<br>Low;<br>High;<br>Very high |
| Type of health facility for hospitalization | Type of health facility where the household member was hospitalized | Nominal:<br>Health center;<br>General referral hospital;<br>Clinic;<br>Other |
| Dependent Variable |  |  |
| Willingness to pay for health care services (WTP) | Whether the respondent is willing to pay for health care services | Nominal:<br>Yes<br>No |
| Amount willing to pay | Amount (in CDF) the respondent is willing to pay for health care services in case of illness | Discrete quantitative:<br>0;<br>1,000-2,500;<br>2,501-7,500;<br>7,501-15,000;<br>15,001-29,000;<br>29,001-58,000;<br>58,001-107,000;<br>107,001-185,000;<br>185,001-231,000;<br>231,001-571,000 |

**Supplementary Table 2:**
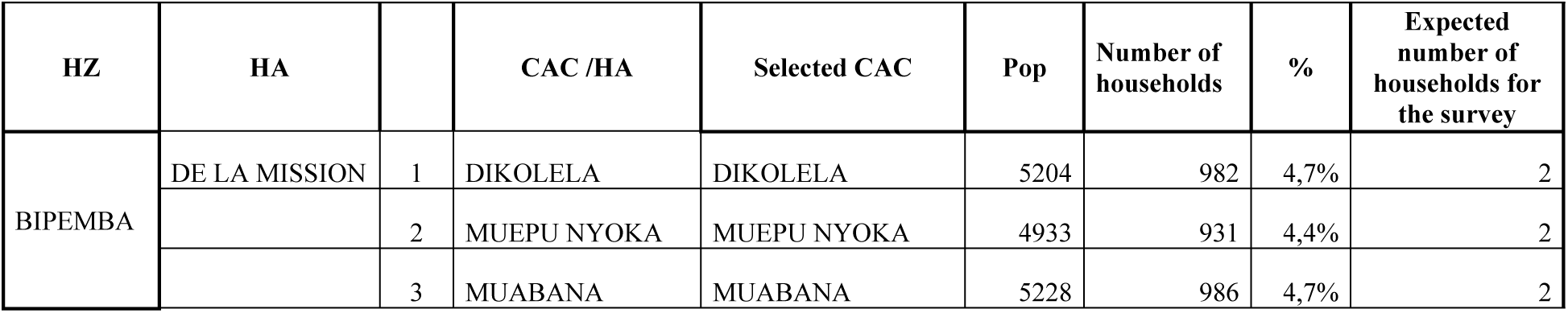
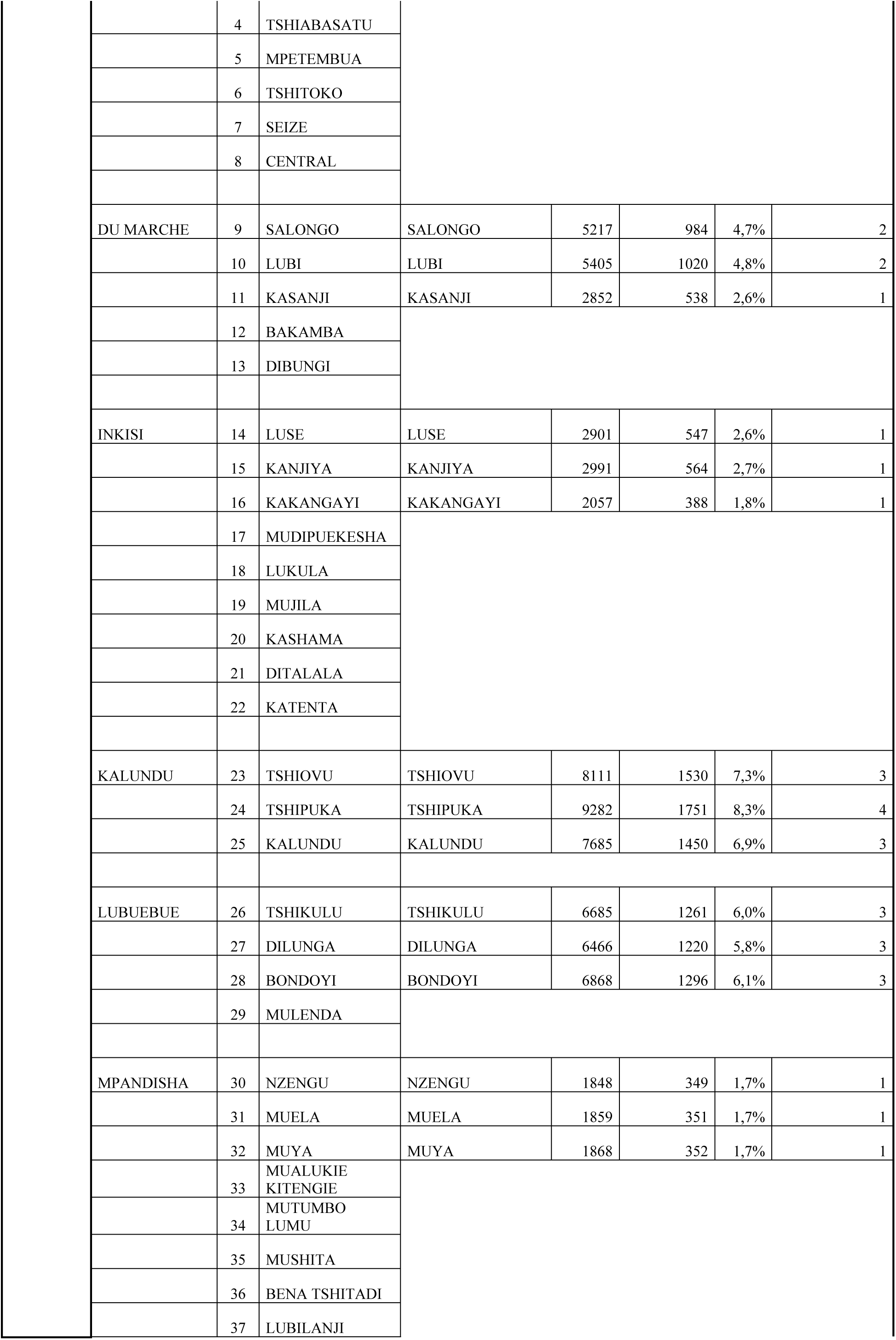
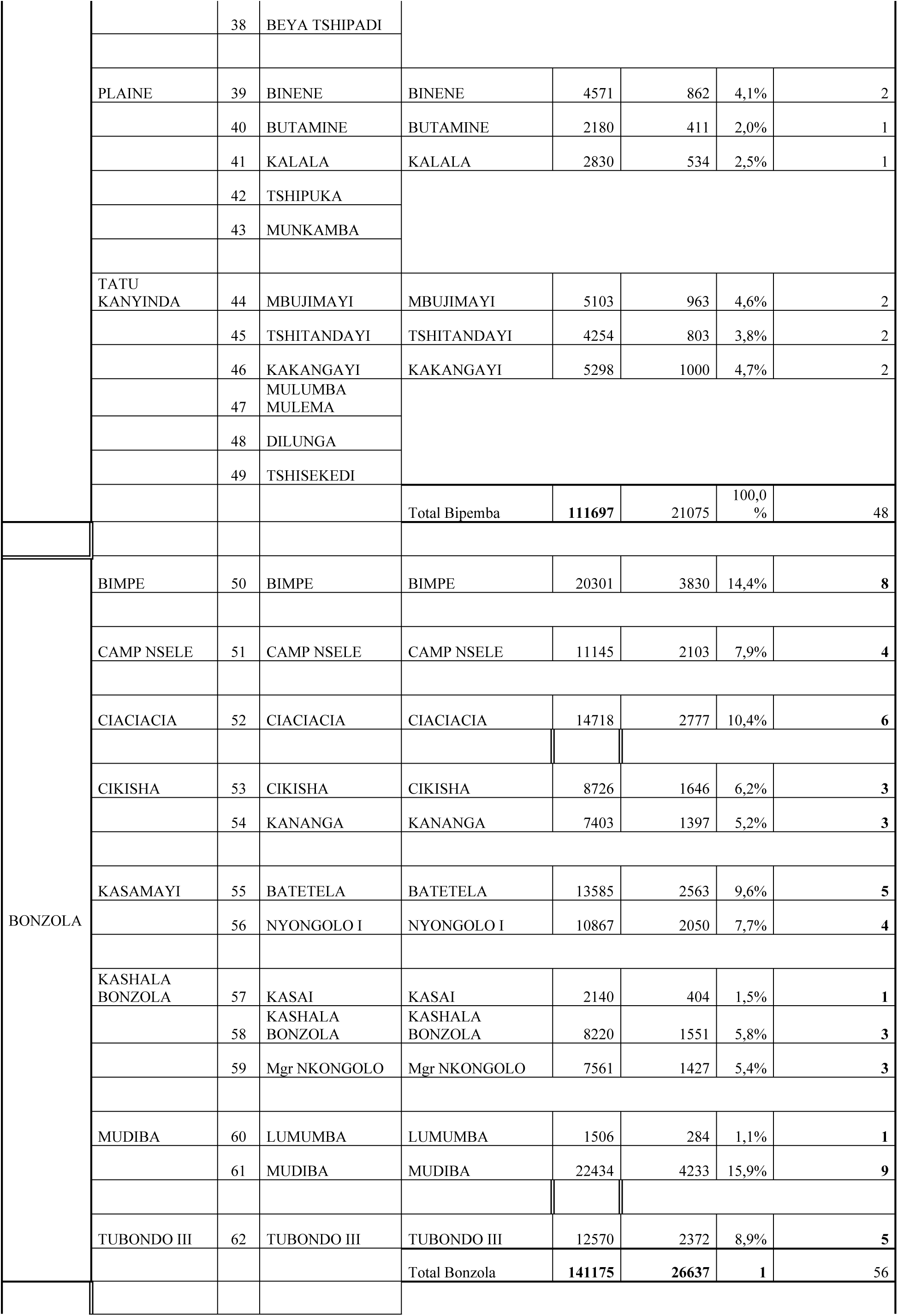
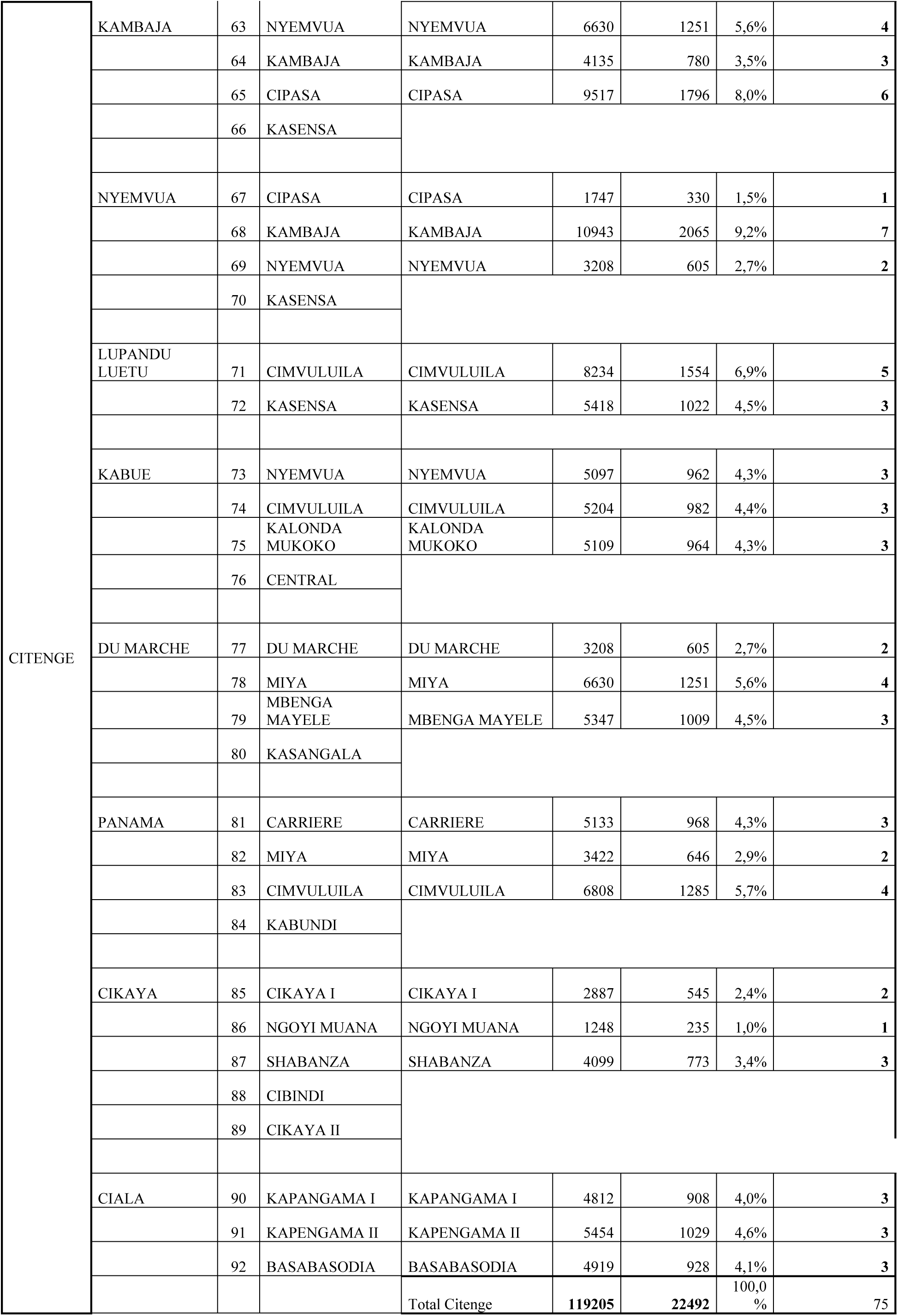
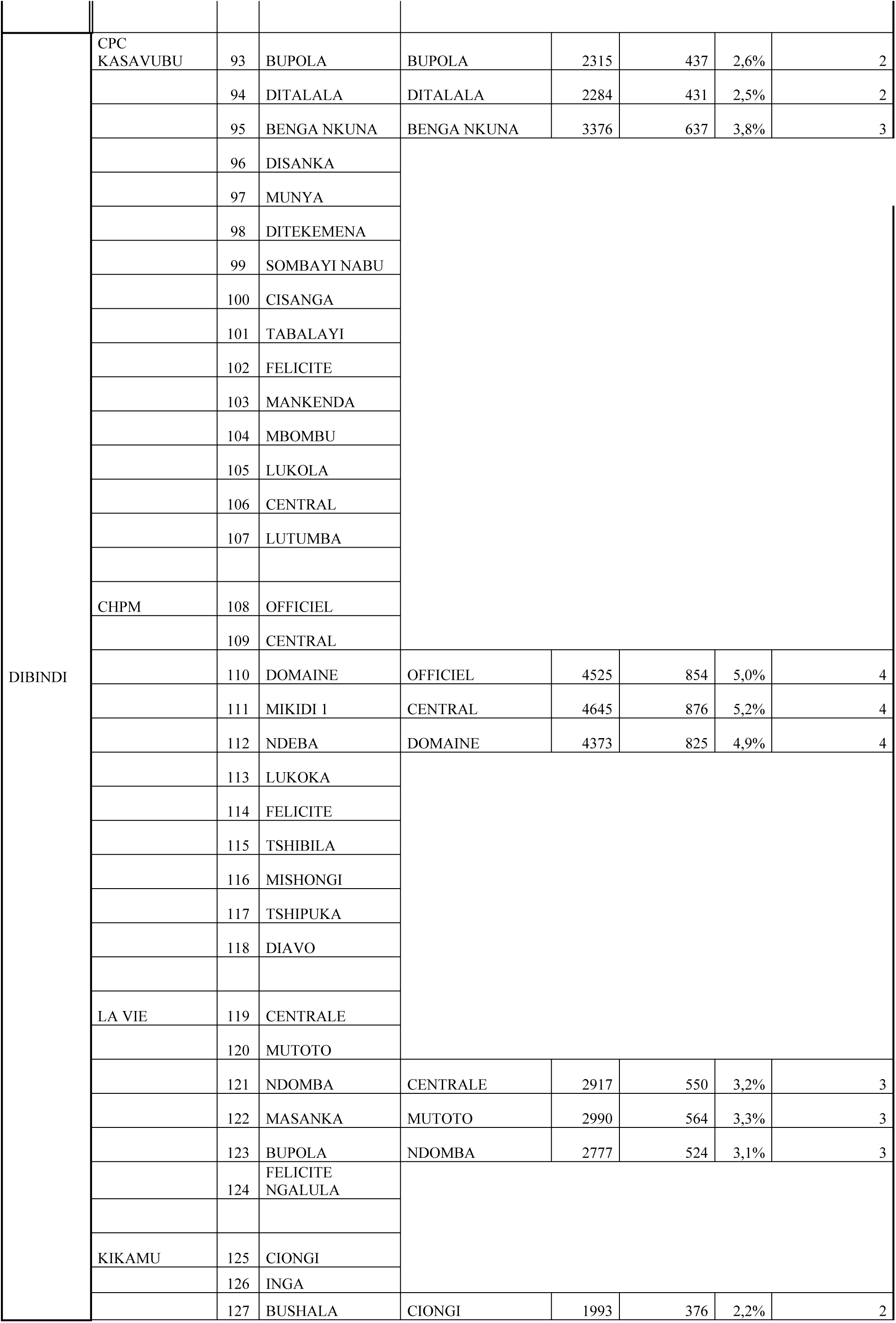
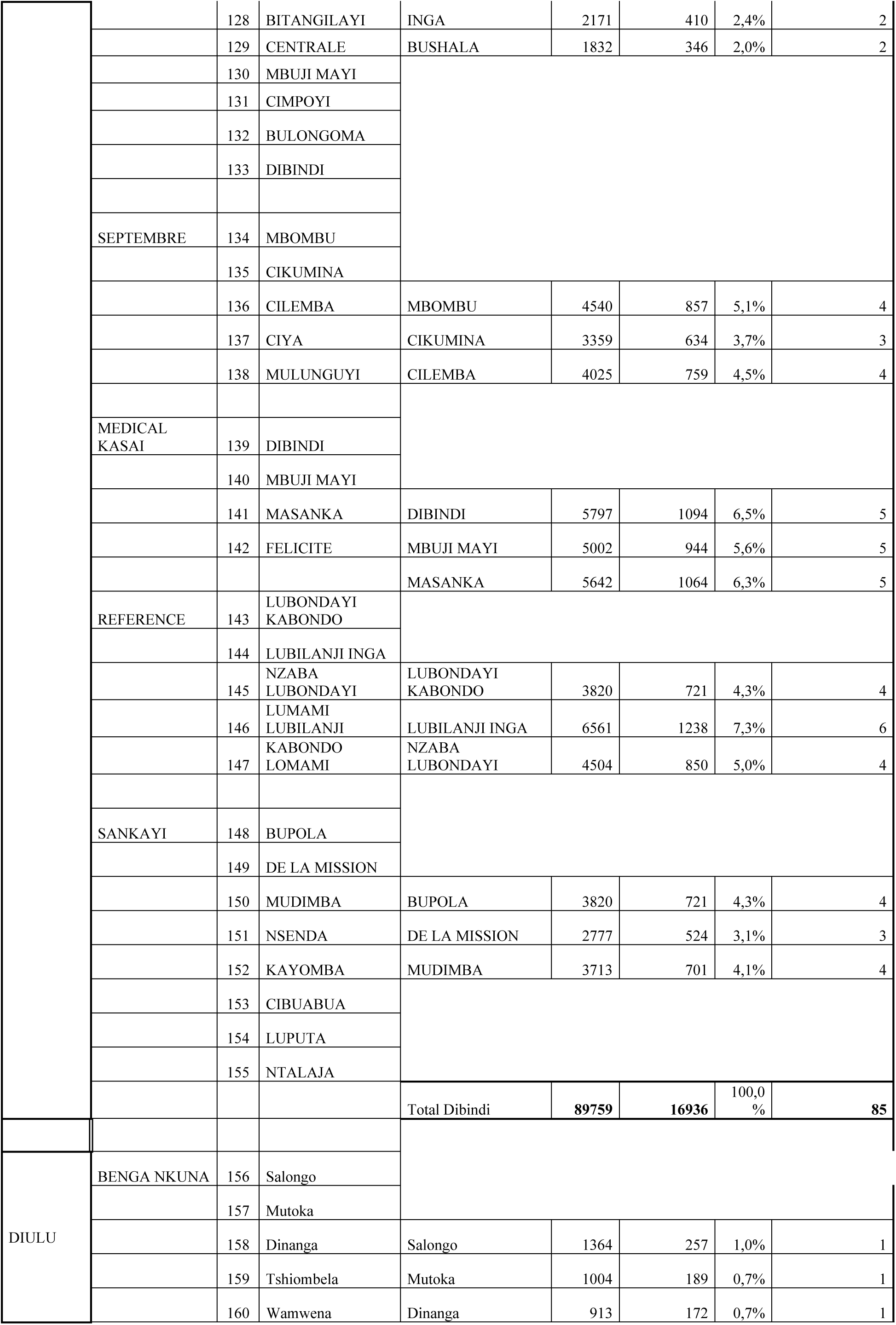
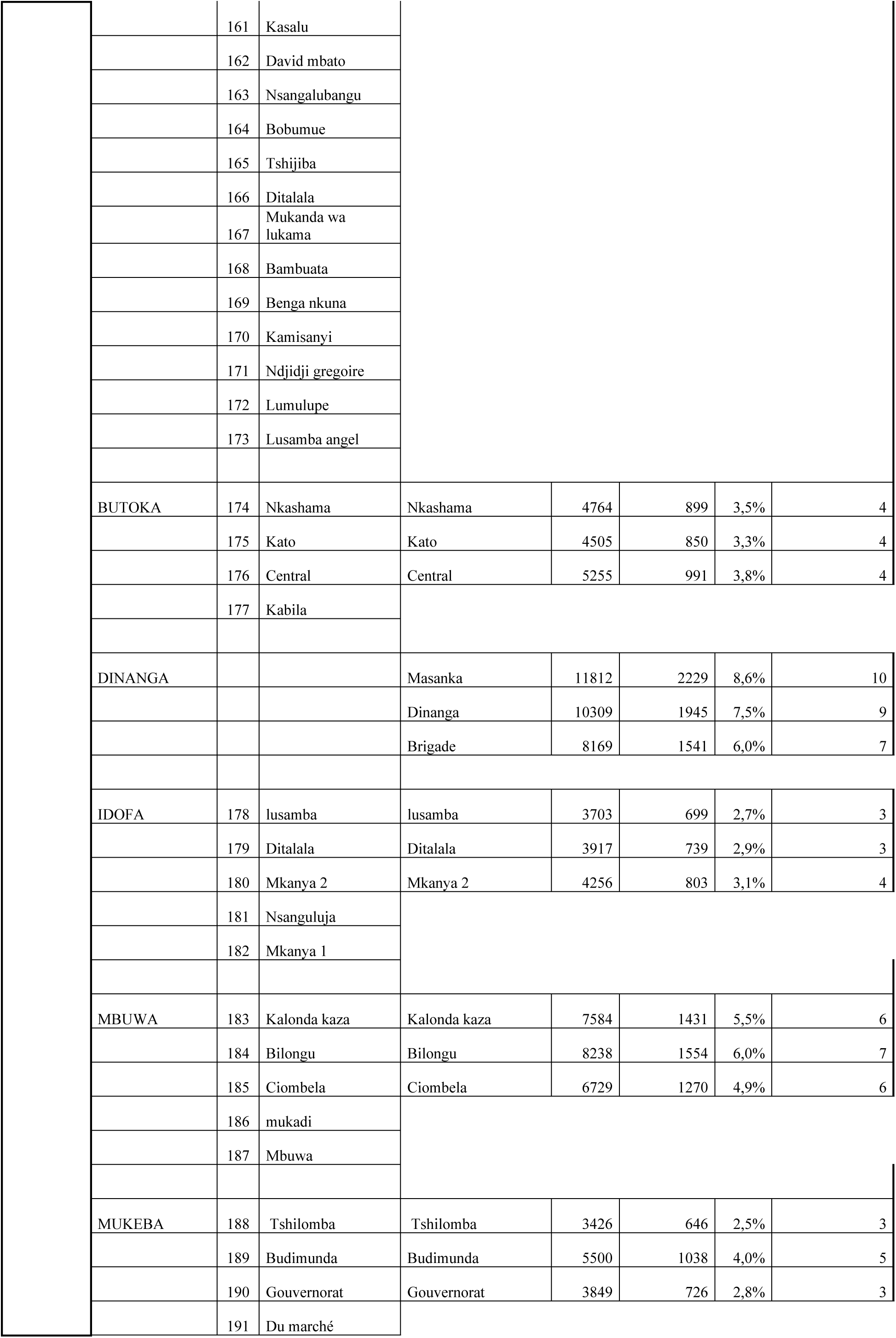
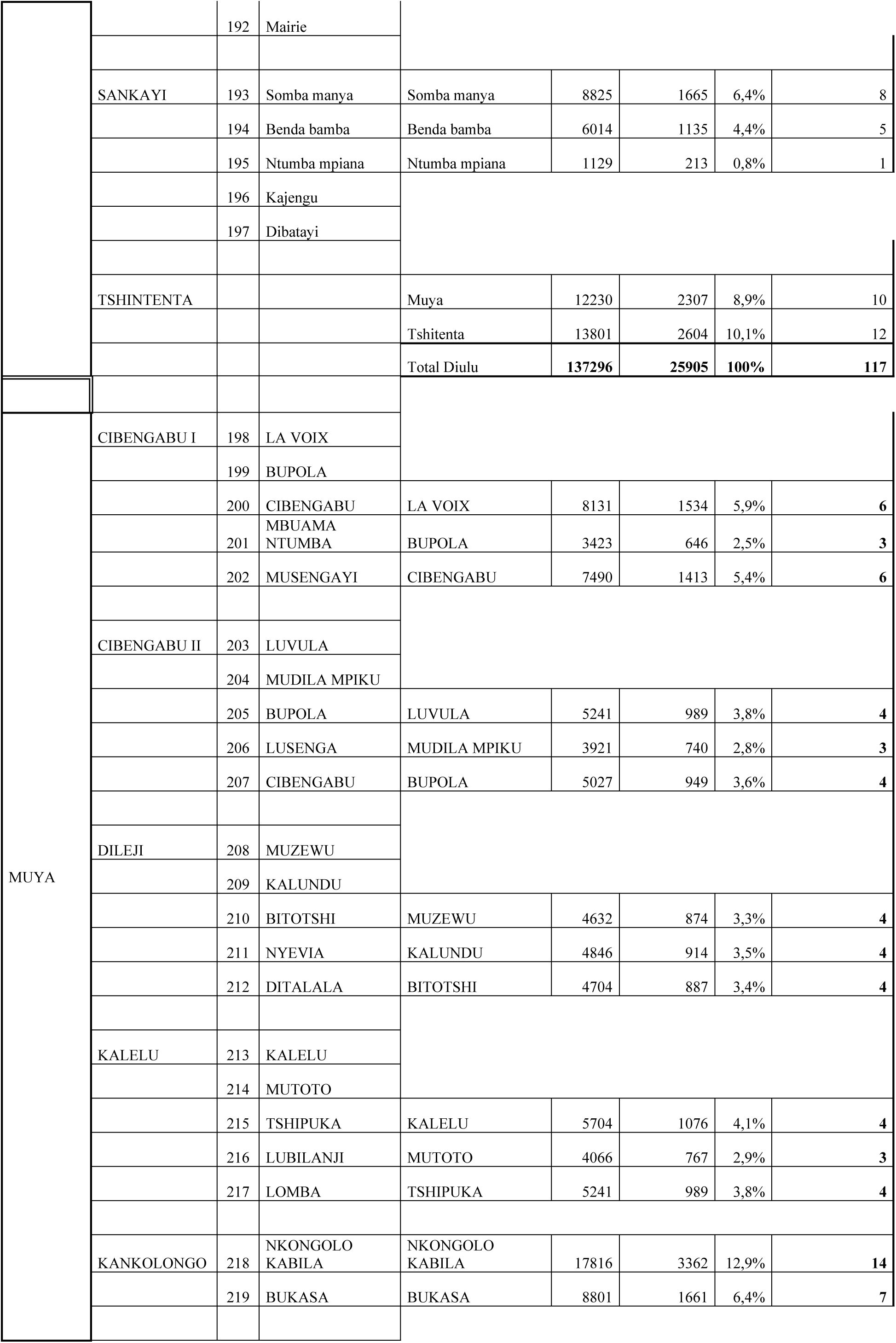
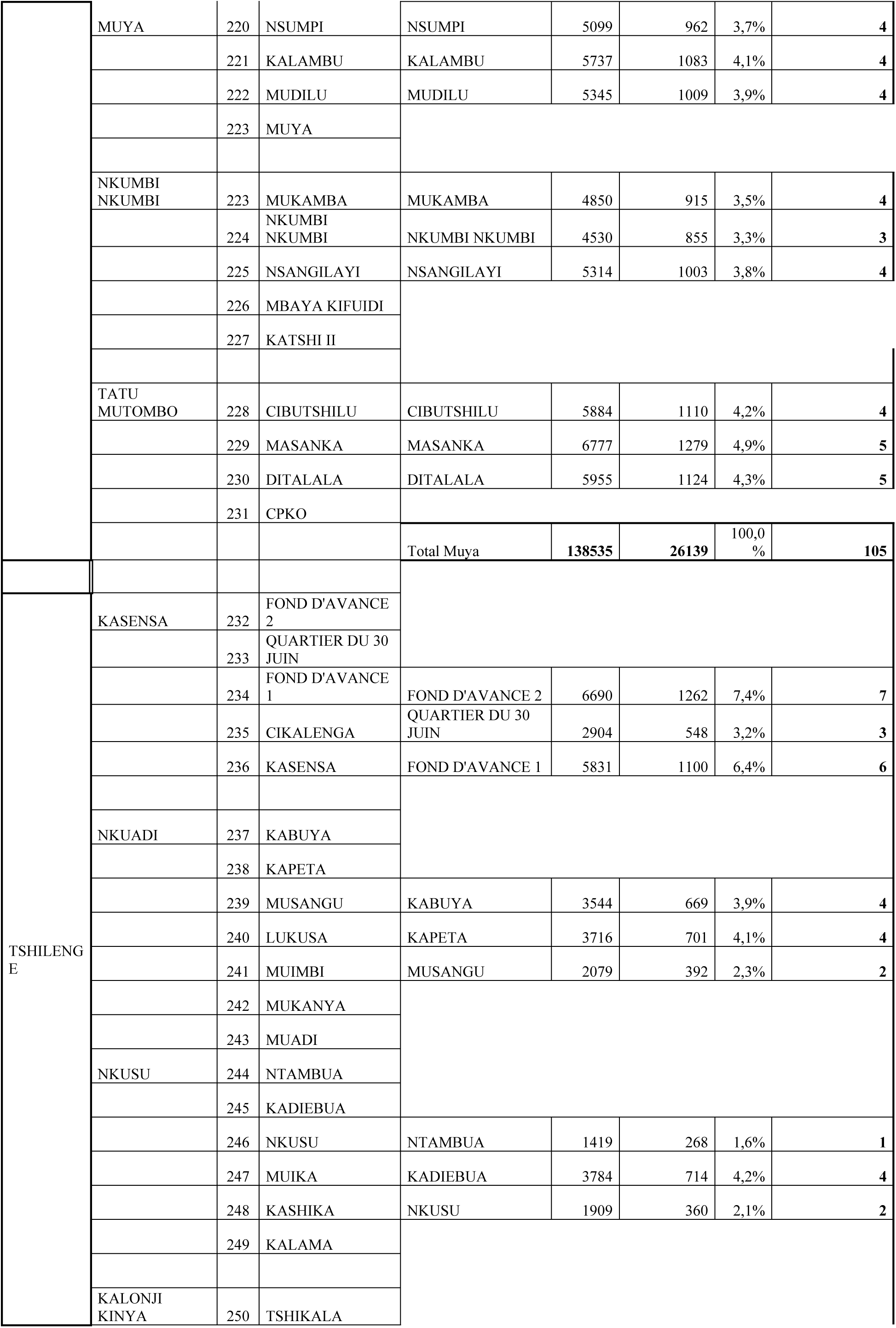
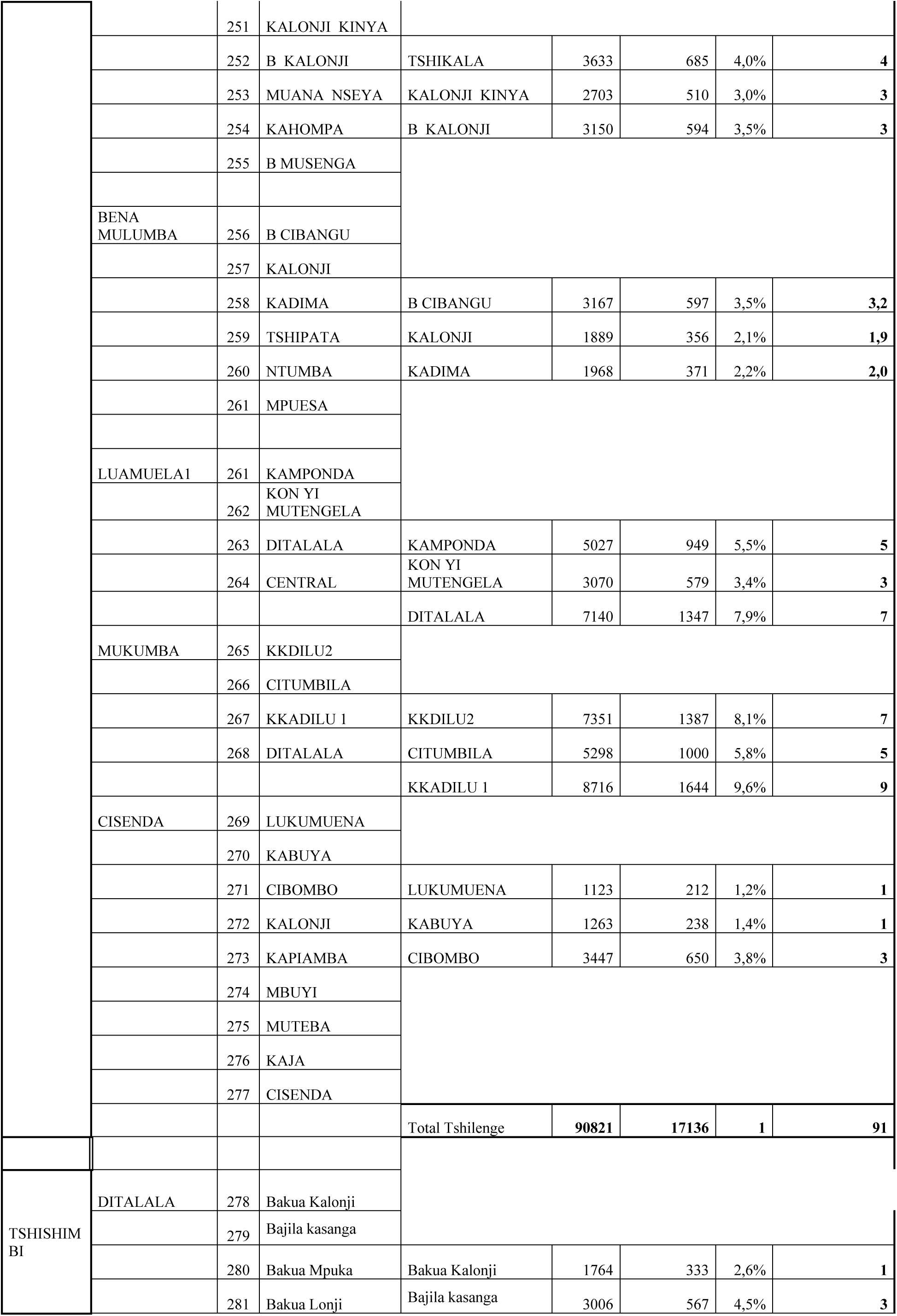
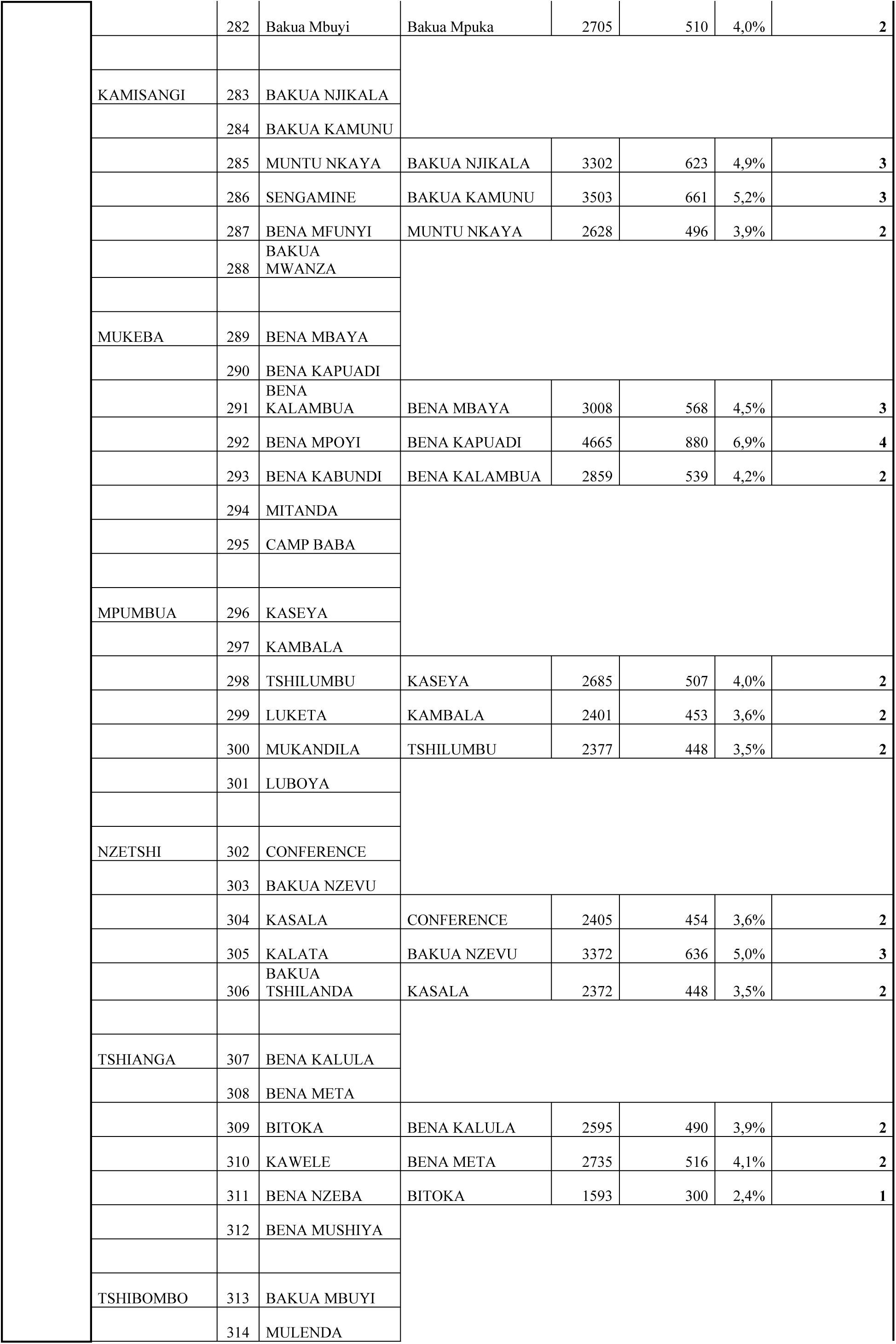
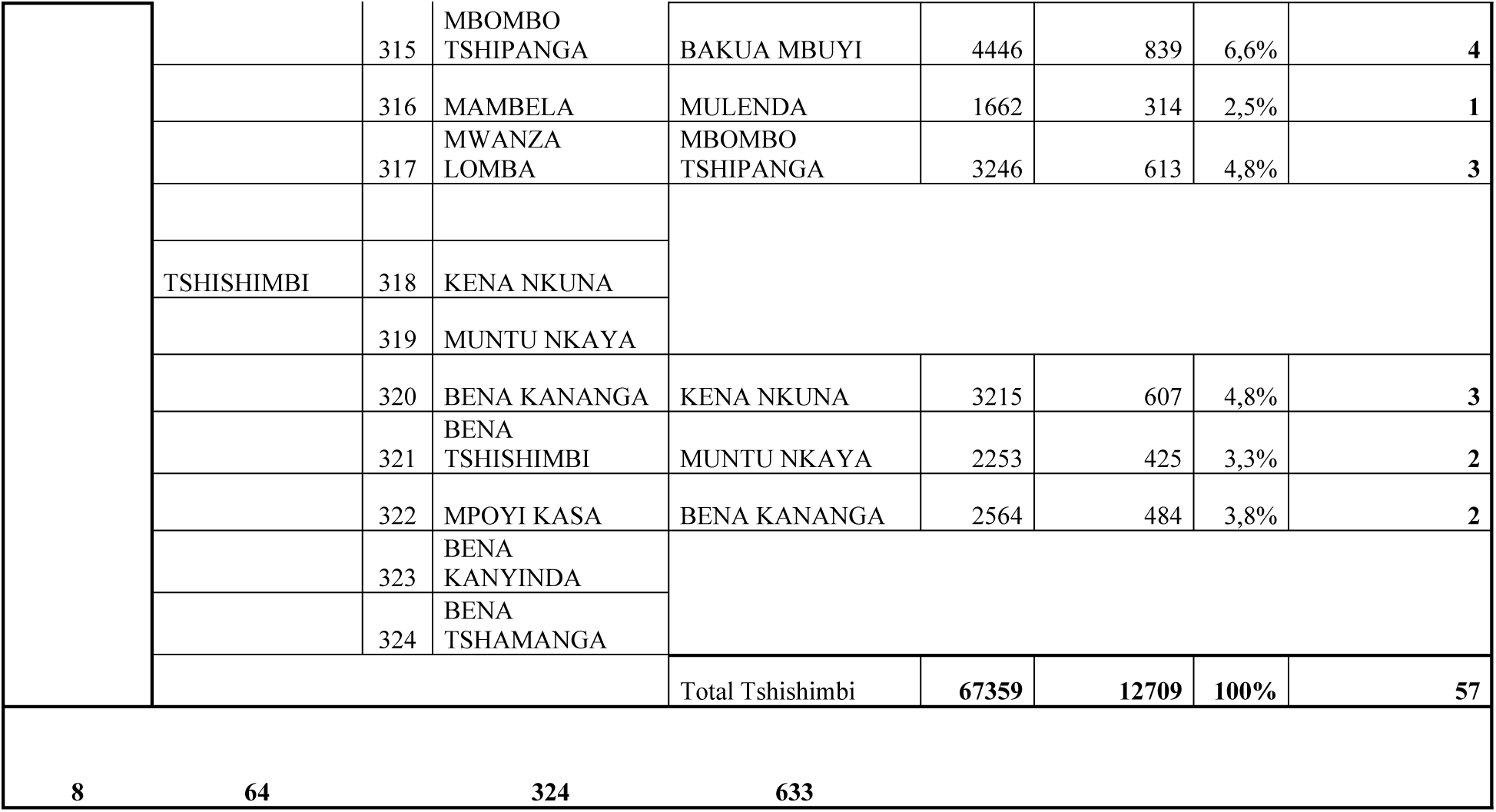
Study sampling design.

## Notes

### Competing Interest Statement

The authors have declared no competing interest.

### Author Declarations

This study was approved by the ethics committee of the University of Mbujimayi under No. 02/CEI/UM/ESU/2025

